# First environmental surveillance for the presence of SARS-CoV-2 RNA in wastewater and river water in Japan

**DOI:** 10.1101/2020.06.04.20122747

**Authors:** Eiji Haramoto, Bikash Malla, Ocean Thakali, Masaaki Kitajima

## Abstract

Wastewater-based epidemiology is a powerful tool to understand the actual incidence of coronavirus disease 2019 (COVID-19) in a community because severe acute respiratory syndrome coronavirus 2 (SARS-CoV-2), the etiological agent of COVID-19, can be shed in the feces of infected individuals regardless of their symptoms. The present study aimed to assess the presence of SARS-CoV-2 RNA in wastewater and river water in Yamanashi Prefecture, Japan, using four quantitative and two nested PCR assays. Influent and secondary-treated (before chlorination) wastewater samples and river water samples were collected five times from a wastewater treatment plant and three times from a river, respectively, between March 17 and May 7, 2020. The wastewater and river water samples (200–5,000 mL) were processed by using two different methods: the electronegative membrane-vortex (EMV) method and the membrane adsorption-direct RNA extraction method. Based on the observed concentrations of indigenous pepper mild mottle virus RNA, the EMV method was found superior to the membrane adsorption-direct RNA extraction method. SARS-CoV-2 RNA was successfully detected in one of five secondary-treated wastewater samples with a concentration of 2.4 × 10^3^ copies/L by N_Sarbeco qPCR assay following the EMV method, whereas all the influent samples were tested negative for SARS-CoV-2 RNA. This result could be attributed to higher limit of detection for influent (4.0 × 10^3^–8.2 × 10^4^ copies/L) with a lower filtration volume (200 mL) compared to that for secondary-treated wastewater (1.4 × 10^2^–2.5 × 10^3^ copies/L) with a higher filtration volume of 5,000 mL. None of the river water samples tested positive for SARS-CoV-2 RNA. Comparison with the reported COVID-19 cases in Yamanashi Prefecture showed that SARS-CoV-2 RNA was detected in the secondary-treated wastewater sample when the cases peaked in the community. This is the first study reporting the detection of SARS-CoV-2 RNA in wastewater in Japan.

## 1. Introduction

Severe acute respiratory syndrome coronavirus 2 (SARS-CoV-2), the etiological agent of the ongoing global pandemic of coronavirus disease 2019 (COVID-19), was first identified in Wuhan, China, in December 2019 (World Health Organization, 2020a). SARS-CoV-2 has spread to 216 countries, areas, or territories with more than 8,006,427 cases and 436,899 deaths worldwide as of June 17, 2020 (World Health Organization, 2020b). A significant amount of infectious SARS-CoV-2 particles were successfully cultured in enterocytes of human small intestinal organoids, where the cellular receptor of SARS-CoV-2, angiotensin-converting enzyme 2 (ACE2), is expressed (Lamers et al., 2020). This report suggests that SARS-CoV-2 actively replicates in enterocytes of human intestine and the virus is subsequently shed in feces. The brush border of intestinal enterocytes is the region where the highest expression of ACE2 can be observed in the human body (Qi et al., 2020). Recent studies reported the detection of SARS-CoV-2 in feces (Wolfel et al., 2020) and urine of COVID-19 patients (Sun et al., 2020; Wang et al., 2020; Wu et al., 2020) at an early onset of infection (Chen et al., 2020; Holshue et al., 2020). A previous study investigating a total of 4,243 COVID-19 patients reported that 17.6% of the patients exhibited gastrointestinal symptoms and SARS-CoV-2 RNA was detected in stool samples from higher proportion (48.1%) of the patients (Cheung et al., 2020). This result indicated that the virus could be shed in feces of infected individuals without gastrointestinal symptoms in addition to patients with diarrhea. The reported proportion of asymptomatic infection of COVID-19 ranges from 18% to 31% (Mizumoto et al., 2020; Nishiura et al., 2020; Treibel et al., 2020), and 21% of COVID-19 patients showed diarrheal symptoms (Wan et al. 2020), which means a large number of symptomatic and asymptomatic individuals shed the virus in stool, which ultimately reaches a wastewater treatment plant (WWTP) through sewage pipes. Even after a patient recover from the respiratory symptoms, viruses can still be shed in feces for several days (Wu et al., 2020).

A recent study reported that SARS-CoV-2 RNA could be detected for longer duration in feces (median, 22 days) than in respiratory airways (18 days) and in serum samples (16 days) (Zheng et al., 2020). While the persistence of SARS-CoV-2 in wastewater has yet not been studied, the time to achieve 99.9% die-off at 23 °C for other coronaviruses (feline infectious peritonitis virus and human coronavirus 229E) was reported to be 2–3 days in sewage (Gundy et al., 2009), while the other coronaviruses persisted for longer duration at low temperature (Casanova et al., 2009). All these pieces of evidence suggested that SARS-CoV-2 can be detected from wastewater and the data can be utilized for wastewater-based epidemiology (WBE). The detection of the SARS-CoV-2 RNA in wastewater in a low COVID-19 prevalent area and detection even before COVID-19 cases was reported by the local authority highlights the importance of wastewater surveillance to monitor the prevalence of the virus (Kitajima et al., 2020; Medema et al., 2020; Randazzo et al., 2020). These recent reports suggested that WBE could provide an early warning sign of possible disease outbreaks in a community (Orive et al., 2020; Xagoraraki and O’Brien, 2020). Such WBE is of particular importance to analyze the data retrospectively in estimating the probable population affected by the virus, because asymptomatic patients who are underdiagnosed by clinical surveillance may also shed the virus in feces.

SARS-CoV-2 RNA has been detected in wastewater in Australia, Italy, Spain, the Netherlands (Ahmed et al., 2020; La Rosa et al., 2020; Medema et al., 2020; Randazzo et al., 2020). The first case of COVID-19 in Japan was reported on January 16, 2020, followed by the first reported case in Yamanashi Prefecture on March 6, 2020. Yamanashi Prefecture is located in the suburbs of Tokyo, and the total cumulative cases of COVID-19 in Tokyo Prefecture reached 5,249 and 64 in Yamanashi Prefecture as of June 1, 2020 (Ministry of Health, Labour and Welfare, 2020). Considering the neighboring prefectures and frequent movement of people between these two prefectures, there is a possibility of the spread of COVID-19 cases in Yamanashi Prefecture, which is currently considered a low prevalence area of COVID-19.

Based on this background, this study aimed to assess the presence of SARS-CoV-2 RNA in wastewater and river water in Yamanashi Prefecture using selected currently available quantitative polymerase chain reaction (qPCR) and nested PCR assays. To the best of our knowledge, this is the first study reporting the detection of SARS-CoV-2 RNA in a water sample in Japan.

## 2. Materials and methods

### 2.1 Collection of water samples

A total of thirteen grab water samples, including five samples each from influent and secondary-treated wastewater before chlorination of a WWTP where conventional activated sludge process was utilized and three river water samples from a river in Yamanashi Prefecture, were collected on five and three different occasions, respectively, between March 17 and May 7, 2020. Secondary-treated wastewater was further treated by chlorination before discharge to the environment; however, the final effluent samples were not collected in this study. Samples were collected in sterilized 1-L plastic bottles taking precautionary measures and transported to the laboratory on ice and processed within 6 h of collection.

### 2.2 Enumeration of Escherichia coli

*E. coli* in wastewater samples was enumerated by a culture-based method using a CHROMagar ECC (Kanto Chemical, Tokyo, Japan), while both the CHROMagar ECC method and a most probable number (MPN) method using a Colilert 18 reagent (IDEXX Laboratories, Westbrook, CA, USA) were used for river water samples, according to the manufacturers’ instructions.

### 2.3 Virus concentration, RNA extraction, and reverse transcription (RT)

Concentration of viruses and RNA extraction were performed by using two methods. In one method, viruses in water samples were concentrated using the electronegative membrane-vortex (EMV) method (Haramoto et al., 2011, 2012) with slight modifications as described previously (Malla et al., 2019). Briefly, 2 mL and 50 mL of 2.5 M MgCl_2_ were added to 200 mL of influent wastewater and 5,000 mL of secondary-treated wastewater samples, respectively, prior to the filtration. For the river water samples, 50 mL of 2.5 M MgCl_2_ was added to 5 L of water samples, which was filtered through a membrane until the membrane clogged. The wastewater and river water samples were filtered through a mixed cellulose-ester membrane (pore size, 0.8 µm; diameter, 90 mm; Merck Millipore, Billerica, MA, USA). Subsequently, 10 mL of an elution buffer containing 0.2 g/L sodium diphosphate decahydrate (Na_4_P_2_O_7_ 10H_2_O), 0.3 g/L ethylenediaminetetraacetic acid trisodium salt trihydrate (C_10_H_13_N_2_O_8_Na_3_ 3H_2_O), and 0.1 mL/L Tween 80 (polyoxyethylene (20) sorbitanmonooleate) was added in a 50-mL plastic tube containing the membrane. The elution step was performed by vigorous vortexing of the membrane with a football-shaped stirring bar. This procedure was repeated using an additional 5 mL of the elution buffer to obtain a final volume of approx. 15 mL. This was followed by a centrifugation step at 2,000 × *g* for 10 min at 4 °C to obtain the supernatant. A disposable membrane filter unit (pore size, 0.45 µm; diameter, 25 mm; Advantec, Tokyo, Japan) was used for the filtration of the supernatant. The filtrate was subsequently concentrated using a Centriprep YM-50 ultrafiltration device (Merck Millipore) to obtain a virus concentrate. One hundred and forty microliters of the virus concentrate was for viral RNA extraction with a QIAamp Viral RNA Mini Kit (Qiagen, Hilden, Germany) in a QIAcube automated platform (Qiagen) to obtain a 60-µL RNA extract. In the second method, which is termed here as the adsorption-direct RNA extraction method, the water samples with 25 mM MgCl_2_ were filtered through a mixed cellulose-ester membrane (pore size, 0.8 µm; diameter, 90 mm; Merck Millipore) and RNA was extracted directly from 1/4 of the membrane that was inserted in a 5-mL PowerWater bead tube of an RNeasy PowerWater Kit (Qiagen). Finally, 50-µL RNA extract was obtained according to the manufacturer’s instruction.

A High-Capacity cDNA Reverse Transcription Kit (Applied Biosystems, Foster City, CA, USA) was used to obtain a 60-µL cDNA from a 30 µL of viral RNA for both methods, following the manufacturer’s protocol.

As recommended previously (Haramoto et al., 2018), coliphage MS2 (ATCC 15597-B1) was added to the sample prior to RNA extraction as a molecular process control. In the case of the EMV method, 1 µL of coliphage MS2 was added to 140 µL each of the virus concentrates and a non-inhibitory control (NIC) sample (PCR-grade water). For the adsorption-direct RNA extraction method, 1 µL of coliphage MS2 was added in a 5-mL PowerWater bead tube containing the filter membrane and 1 mL of the Solution PM1 from the RNeasy PowerWater kit (Qiagen) and β-mercaptoethanol. The concentration of coliphage MS2 in a sample and NIC tubes were determined using qPCR (Friedman et al., 2011) and the extraction-RT-qPCR efficiencies were calculated as the ratio of the concentration of cDNA in a sample qPCR tube to that in an NIC tube.

The calculated extraction-RT-qPCR efficiencies were 71.6 ± 25.2% (n = 13) and 8.5 ± 3.7% (n = 11) for the EMV and the adsorption-direct RNA extraction methods, respectively, indicating that there was no substantial loss and/or inhibition in the water samples during RNA extraction, RT, and qPCR. The resultant low extraction-RT-qPCR efficiency of the adsorption-direct RNA extraction method could be due to an assumption that there was a 100% recovery of virus by the EMV method.

### 2.4 qPCR and nested PCR assays

As shown in Tables 1 and 2, a total of six recently published assays, including four qPCR assays (N_Sarbeco, NIID_2019-nCOV_N, and CDC-N1 and -N2 assays) (Centers for Disease Control and Prevention, 2020; Corman et al., 2020; Shirato et al., 2020) and two nested PCR assays (ORF1a and S protein assays) (Shirato et al., 2020), were applied for the detection of SARS-CoV-2 RNA in the wastewater and river water samples. In addition, an RT-qPCR assay targeting pepper mild mottle virus (PMMoV), a plant virus originating from pepper products, which is considered as a potential viral indicator of human fecal contamination, was tested (Rosario et al., 2009; Hamza et al., 2011; Kuroda et al., 2014; Kitajima et al., 2018; Symonds et al., 2018).

**Table 1.**
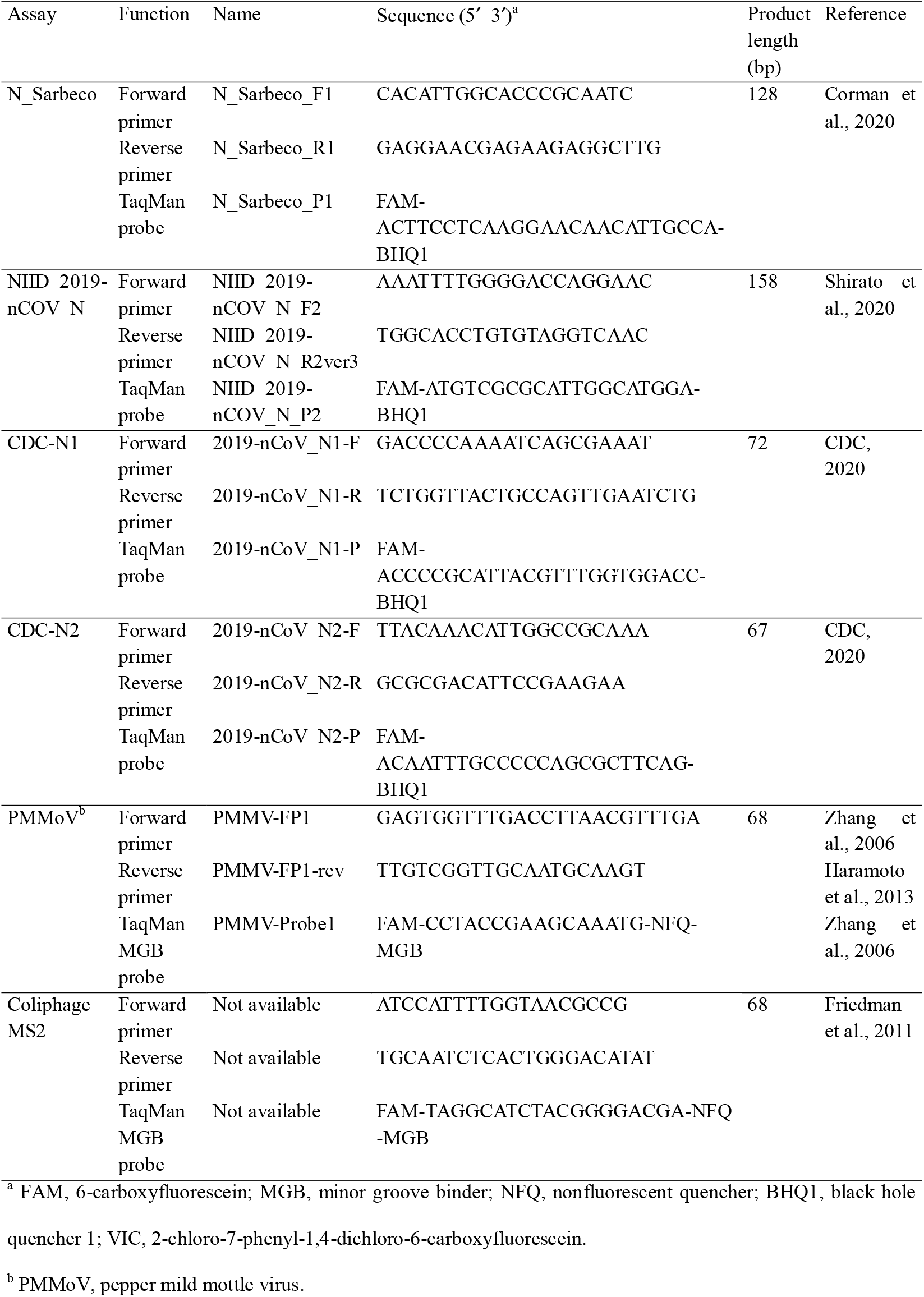
Primers and probes of qPCR assays used in this study

**Table 2.**
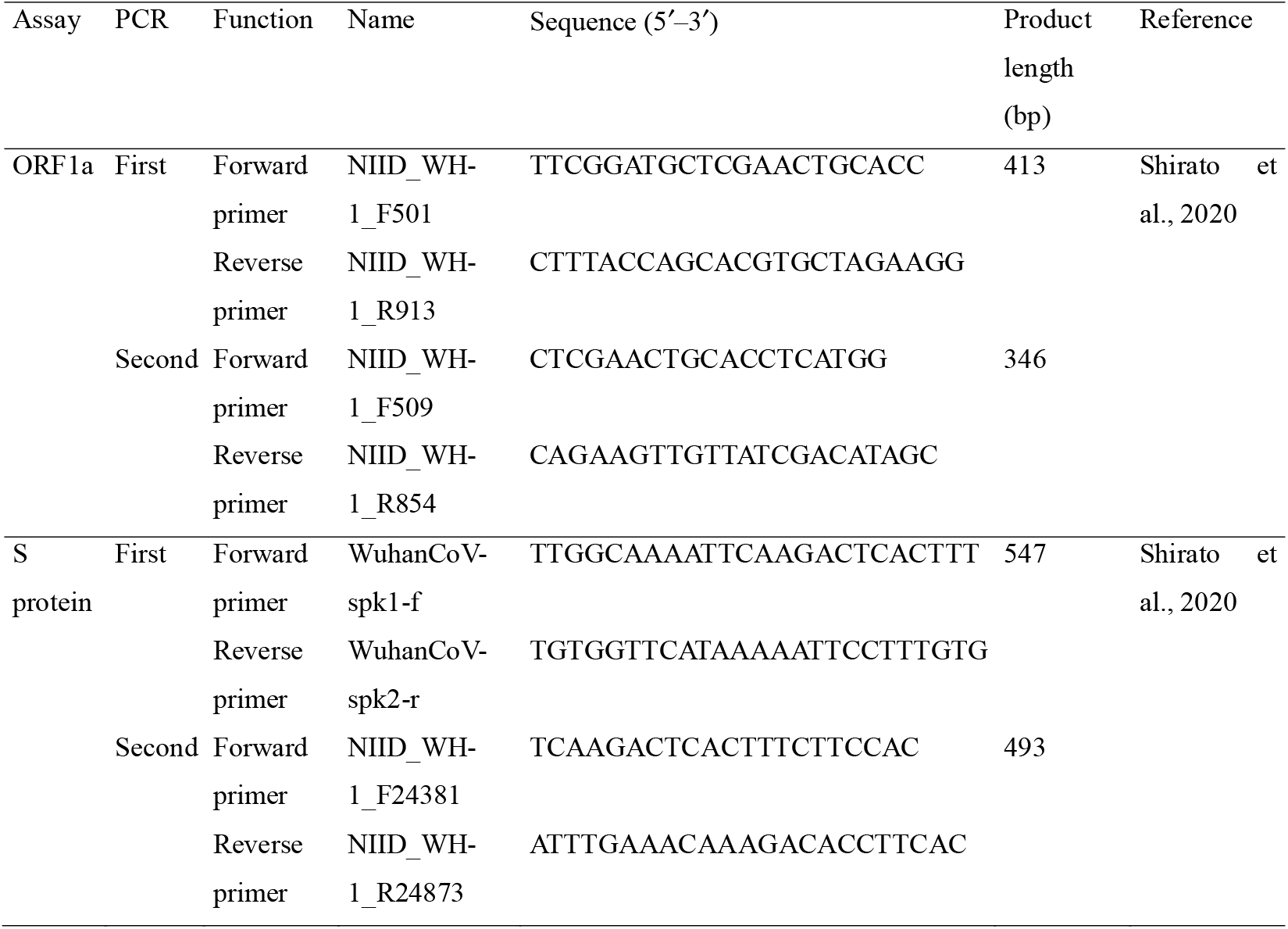
Primers and probes of nested PCR assays used in this study

RT-qPCR assays for SARS-CoV-2 were performed in 25-µL qPCR reaction volume containing 12.5 µL of Probe qPCR Mix with UNG (Takara Bio, Kusatsu, Japan), 0.1 µL each of 100 µM forward and reverse primers, 0.05 µL of 100 µM TaqMan probe, and 2.5 µL of template cDNA. Nested PCR assays for SARS-CoV-2 were performed in a 50-µL reaction mixture. First PCR amplification reaction mixture contained 25 µL of *Premix Ex Taq* Hot Start Version (Takara Bio), 0.15 µL each of 100 µM forward and reverse primers, and 5.0 µL of template cDNA, while in second PCR amplification, 2 µL of first PCR product was used in a total volume of 50 µL. The thermal cycling conditions of the qPCR assays were as follows: initial incubation at 25 °C for 10 min and initial denaturation at 95 °C for 30 s, followed by 45 cycles of denaturation at 95 °C for 5 s and primer annealing and extension reaction at 60 °C for 60 s (for N_Sarbeco, NIID_2019-nCOV_N, and PMMoV), or at 60 °C for 30 s (for CDC-N1 and -N2). The thermal cycling conditions for both first and second rounds of nested PCR assays were as follows: initial denaturation at 95 °C for 2 min, followed by 35 cycles of denaturation at 94 °C for 30 s and primer annealing at 56 °C for 30 s, and extension reaction at 72 °C for 60 s, and a final extension at 72 °C for 7 min.

A Thermal Cycler Dice Real Time System TP800 (Takara Bio) was used for the quantification for the qPCR assays, while gel electrophoresis was performed in 2% agarose gel to visualize the nested PCR products under ultraviolet light. Five to six 10-fold serial dilutions of gBlocks gene fragments containing the amplification region sequences of the qPCR assays (Integrated DNA Technologies, Coralville, IA, USA) were used to generate a standard curve. Negative and positive controls were included in every qPCR run, and all samples were tested with duplicated qPCR reactions. For the nested PCR assays, one negative and one positive control (gBlocks) was included in each gel electrophoresis run.

### 2.5 Statistical analysis

The paired *t*-test was used to compare the performance of the two different concentration-RNA extraction methods. One-tenth of the lower limit of detection (LOD) value of PMMoV (1.0 × 10^1^ copies/L) was used for the negative sample for the statistical analysis, as described previously (Malla et al., 2018). Microsoft Office Excel 2013 (Microsoft Corporation, Redmond, WA, USA) was used to perform the statistical analysis, and a significant value was set at 0.05.

## 3. Results

### 3.1 Detection of E. coli and PMMoV in wastewater and river water samples

The influent and secondary-treated wastewater samples (n = 5 each) were tested to determine the reduction efficiency of *E. coli*, a conventional fecal indicator bacterium, and PMMoV, the most abundant virus in wastewater (Kitajima et al., 2014) and proposed as an indicator of virus reduction (Kitajima et al., 2018; Symonds et al., 2018; Tandukar et al., 2020). The geometric mean concentrations of *E. coli* and PMMoV RNA in the influent samples were 3.7 × 10^4^ colony forming-units (CFU)/mL (range, 1.8 × 10^4^–6.7 × 10^4^CFU/mL; n = 5) and 4.8 × 10^7^ copies/L (range, 3.2 × 10^7^–9.4 × 10^7^ copies/L; n = 5), respectively. The log_10_ reduction ratios (mean ± standard deviation (SD)) of *E. coli* and PMMoV RNA based on their concentrations in influent and secondary-treated wastewater samples were 2.7 ± 0.3 log_10_ (n = 5) and 1.8 ± 0.2 log_10_ (n = 5), respectively. *E. coli* was detected in all river water samples (n = 3) by the CHROMagar ECC method and the MPN method with geometric mean concentrations of 1.2 CFU/mL (range, 0.2–5.0 CFU/mL) and 3.8 MPN/100 mL (range, 9.1 × 10^1^−1.2 × 10^3^ MPN/100 mL), respectively. PMMoV RNA was detected with a geometric mean concentration of 2.7 × 10^5^ copies/L (range, 1.8 × 10^5^−4.3 × 10^5^ copies/L; n = 3) in the river water samples.

### 3.2 Comparison of concentration-RNA extraction methods

Table 3 summarizes the comparison of the two concentration-RNA extraction methods, based on the observed concentrations of indigenous PMMoV RNA. Using the EMV method, PMMoV RNA was detected in 100% (13/13) of wastewater and river water samples, while using an adsorption-direct RNA extraction method, it was detected in 91% (10/11) of the samples. The geometric mean concentration of PMMoV RNA using the EMV method (2.6 × 10^6^ copies/L; range, 1.8 × 10^5^−1.0 × 10^8^ copies/L; n = 11) was significantly higher than that using the adsorption-direct RNA extraction method (1.3 × 10^4^ copies/L; range, < 1.0 × 10^1^− 1.3 × 10^6^ copies/L; n = 11) (paired *t*-test; *p* < 0.05). Assuming the concentration-RNA extraction-RT-qPCR efficiency of PMMoV by the EMV method as 100%, the efficiency by adsorption-direct RNA extraction method was 1.5 ± 2.3% (n = 11).

**Table 3.** Comparison of virus concentration-RNA extraction methods

| Sample ID | Date of sample collection<br>(mm/dd/yyyy) | PMMoV (copies/L) |  |
| --- | --- | --- | --- |
|  |  | EMV method<sup>a</sup> | Adsorption-direct RNA extraction method<sup>b</sup> |
| Influent | 4/14/2020 | $3.2 \times 10^7$ | $2.7 \times 10^5$ |
| | 4/22/2020 | $5.0 \times 10^7$ | $5.3 \times 10^5$ |
| | 4/30/2020 | $4.8 \times 10^7$ | $7.0 \times 10^5$ |
| | 5/7/2020 | $1.0 \times 10^8$ | $1.3 \times 10^6$ |
| Secondary-treated wastewater | 4/14/2020 | $3.8 \times 10^5$ | $3.1 \times 10^4$ |
| | 4/22/2020 | $1.2 \times 10^6$ | $7.0 \times 10^3$ |
| | 4/30/2020 | $4.8 \times 10^5$ | $1.3 \times 10^4$ |
| | 5/7/2020 | $1.3 \times 10^6$ | $1.6 \times 10^3$ |
| River water | 4/22/2020 | $1.8 \times 10^5$ | $<1.0 \times 10^1$ |
| | 4/30/2020 | $2.6 \times 10^5$ | $1.2 \times 10^2$ |
| | 5/7/2020 | $4.0 \times 10^5$ | $2.5 \times 10^3$ |

<sup>a</sup> Water sample concentrated by the electronegative membrane-vortex (EMV) method, followed by RNA extraction using the QIAamp Viral RNA Mini Kit (Qiagen); <sup>b</sup> Pretreated water samples filtered using a mixed cellulose-ester membrane and RNA was extracted directly from the filter membrane using the RNeasy PowerWater Kit (Qiagen).

### 3.3 Detection of SARS-CoV-2 RNA in wastewater and river water samples

The collected wastewater (n = 10) and river water samples (n = 3) were processed with both the EMV and the adsorption-direct RNA extraction methods and tested for SARS-CoV-2 RNA using six qPCR/nested PCR assays. As summarized in Figure 1 and Table 4, SARS-CoV-2 RNA was detected in one (20%) secondary-treated wastewater sample, which had been collected on April 14, 2020 and concentrated by the EMV method, using the N_Sarbeco qPCR assay. Meanwhile, SARS-CoV-2 RNA was not detected in any of the influent (n = 5) and river water samples (n = 3) using six qPCR/nested PCR assays by the EMV and the adsorption-direct RNA extraction methods.

**Table 4.**
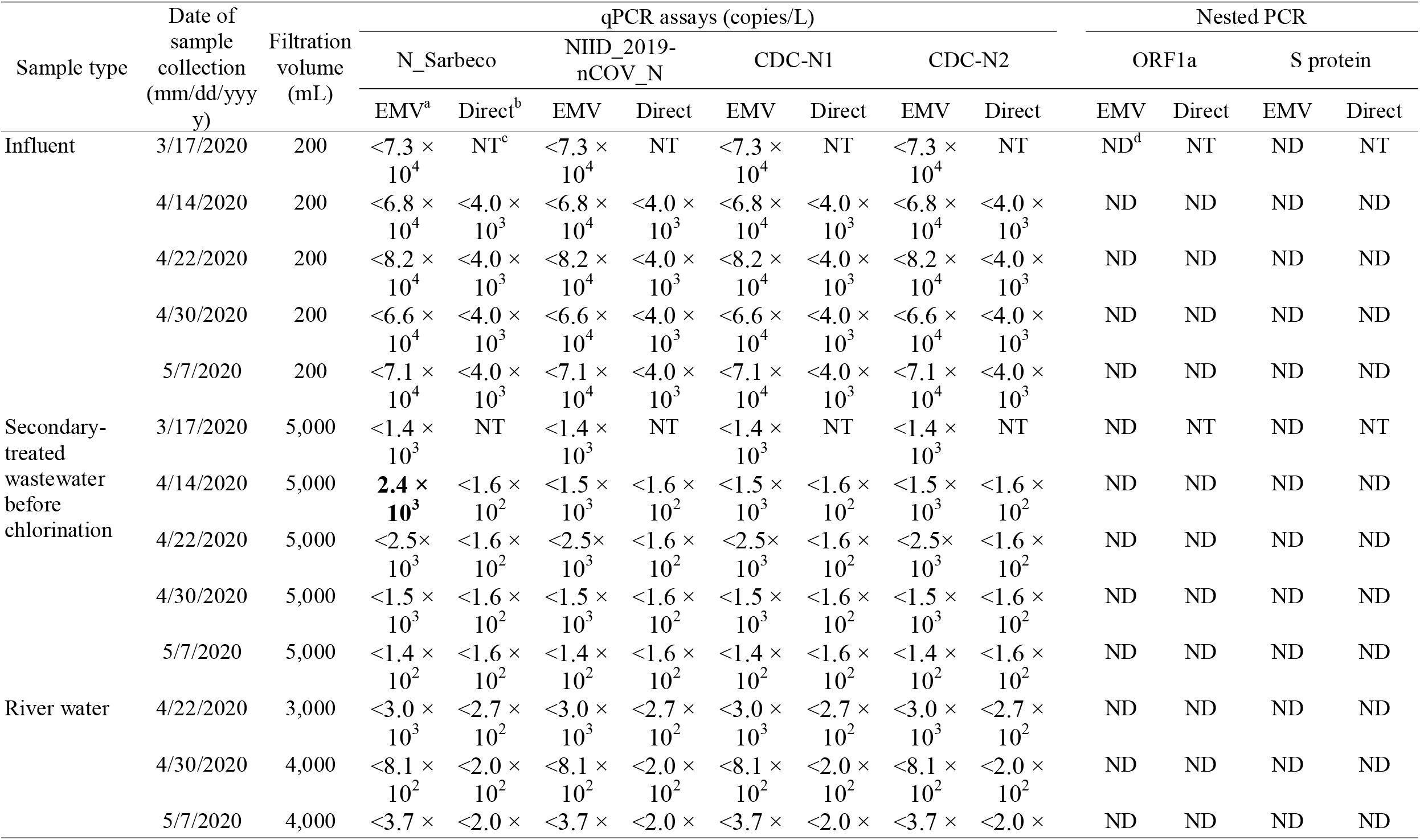
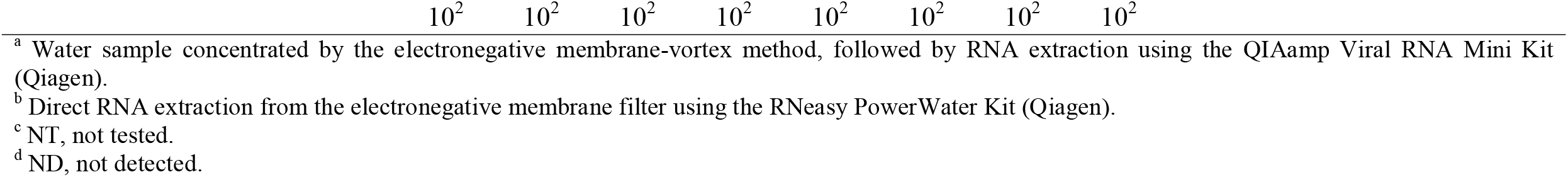
Detection of SARS-CoV-2 RNA in wastewater and river water samples

| Sample type | Date of sample collection (mm/dd/yyyy) | Filtration volume (mL) | qPCR assays (copies/L) |  |  |  |  |  |  |  | Nested PCR |  |  |  |
| --- | --- | --- | --- | --- | --- | --- | --- | --- | --- | --- | --- | --- | --- | --- |
|  |  |  | N_Sarbeco |  | NIID_2019-nCoV_N |  | CDC-N1 |  | CDC-N2 |  | ORF1a |  | S protein |  |
|  |  |  | EMV<sup>a</sup> | Direct<sup>b</sup> | EMV | Direct | EMV | Direct | EMV | Direct | EMV | Direct | EMV | Direct |
| Influent | 3/17/2020 | 200 | <7.3 × 10<sup>4</sup> | NT<sup>c</sup> | <7.3 × 10<sup>4</sup> | NT | <7.3 × 10<sup>4</sup> | NT | <7.3 × 10<sup>4</sup> | NT | ND<sup>d</sup> | NT | ND | NT |
|  | 4/14/2020 | 200 | <6.8 × 10<sup>4</sup> | <4.0 × 10<sup>3</sup> | <6.8 × 10<sup>4</sup> | <4.0 × 10<sup>3</sup> | <6.8 × 10<sup>4</sup> | <4.0 × 10<sup>3</sup> | <6.8 × 10<sup>4</sup> | <4.0 × 10<sup>3</sup> | ND | ND | ND | ND |
|  | 4/22/2020 | 200 | <8.2 × 10<sup>4</sup> | <4.0 × 10<sup>3</sup> | <8.2 × 10<sup>4</sup> | <4.0 × 10<sup>3</sup> | <8.2 × 10<sup>4</sup> | <4.0 × 10<sup>3</sup> | <8.2 × 10<sup>4</sup> | <4.0 × 10<sup>3</sup> | ND | ND | ND | ND |
|  | 4/30/2020 | 200 | <6.6 × 10<sup>4</sup> | <4.0 × 10<sup>3</sup> | <6.6 × 10<sup>4</sup> | <4.0 × 10<sup>3</sup> | <6.6 × 10<sup>4</sup> | <4.0 × 10<sup>3</sup> | <6.6 × 10<sup>4</sup> | <4.0 × 10<sup>3</sup> | ND | ND | ND | ND |
|  | 5/7/2020 | 200 | <7.1 × 10<sup>4</sup> | <4.0 × 10<sup>3</sup> | <7.1 × 10<sup>4</sup> | <4.0 × 10<sup>3</sup> | <7.1 × 10<sup>4</sup> | <4.0 × 10<sup>3</sup> | <7.1 × 10<sup>4</sup> | <4.0 × 10<sup>3</sup> | ND | ND | ND | ND |
| Secondary-treated wastewater before chlorination | 3/17/2020 | 5,000 | <1.4 × 10<sup>3</sup> | NT | <1.4 × 10<sup>3</sup> | NT | <1.4 × 10<sup>3</sup> | NT | <1.4 × 10<sup>3</sup> | NT | ND | NT | ND | NT |
|  | 4/14/2020 | 5,000 | **2.4** × 10<sup>3</sup> | <1.6 × 10<sup>2</sup> | <1.5 × 10<sup>3</sup> | <1.6 × 10<sup>2</sup> | <1.5 × 10<sup>3</sup> | <1.6 × 10<sup>2</sup> | <1.5 × 10<sup>3</sup> | <1.6 × 10<sup>2</sup> | ND | ND | ND | ND |
|  | 4/22/2020 | 5,000 | <2.5 × 10<sup>3</sup> | <1.6 × 10<sup>2</sup> | <2.5 × 10<sup>3</sup> | <1.6 × 10<sup>2</sup> | <2.5 × 10<sup>3</sup> | <1.6 × 10<sup>2</sup> | <2.5 × 10<sup>3</sup> | <1.6 × 10<sup>2</sup> | ND | ND | ND | ND |
|  | 4/30/2020 | 5,000 | <1.5 × 10<sup>3</sup> | <1.6 × 10<sup>2</sup> | <1.5 × 10<sup>3</sup> | <1.6 × 10<sup>2</sup> | <1.5 × 10<sup>3</sup> | <1.6 × 10<sup>2</sup> | <1.5 × 10<sup>3</sup> | <1.6 × 10<sup>2</sup> | ND | ND | ND | ND |
|  | 5/7/2020 | 5,000 | <1.4 × 10<sup>2</sup> | <1.6 × 10<sup>2</sup> | <1.4 × 10<sup>2</sup> | <1.6 × 10<sup>2</sup> | <1.4 × 10<sup>2</sup> | <1.6 × 10<sup>2</sup> | <1.4 × 10<sup>2</sup> | <1.6 × 10<sup>2</sup> | ND | ND | ND | ND |
| River water | 4/22/2020 | 3,000 | <3.0 × 10<sup>3</sup> | <2.7 × 10<sup>2</sup> | <3.0 × 10<sup>3</sup> | <2.7 × 10<sup>2</sup> | <3.0 × 10<sup>3</sup> | <2.7 × 10<sup>2</sup> | <3.0 × 10<sup>3</sup> | <2.7 × 10<sup>2</sup> | ND | ND | ND | ND |
|  | 4/30/2020 | 4,000 | <8.1 × 10<sup>2</sup> | <2.0 × 10<sup>2</sup> | <8.1 × 10<sup>2</sup> | <2.0 × 10<sup>2</sup> | <8.1 × 10<sup>2</sup> | <2.0 × 10<sup>2</sup> | <8.1 × 10<sup>2</sup> | <2.0 × 10<sup>2</sup> | ND | ND | ND | ND |
|  | 5/7/2020 | 4,000 | <3.7 × 10<sup>2</sup> | <2.0 × 10<sup>2</sup> | <3.7 × 10<sup>2</sup> | <2.0 × 10<sup>2</sup> | <3.7 × 10<sup>2</sup> | <2.0 × 10<sup>2</sup> | <3.7 × 10<sup>2</sup> | <2.0 × 10<sup>2</sup> | ND | ND | ND | ND |

<sup>a</sup> Water sample concentrated by the electronegative membrane-vortex method, followed by RNA extraction using the QIAamp Viral RNA Mini Kit (Qiagen).
<sup>b</sup> Direct RNA extraction from the electronegative membrane filter using the RNeasy PowerWater Kit (Qiagen).
<sup>c</sup> NT, not tested.
<sup>d</sup> ND, not detected.

**Fig. 1.**
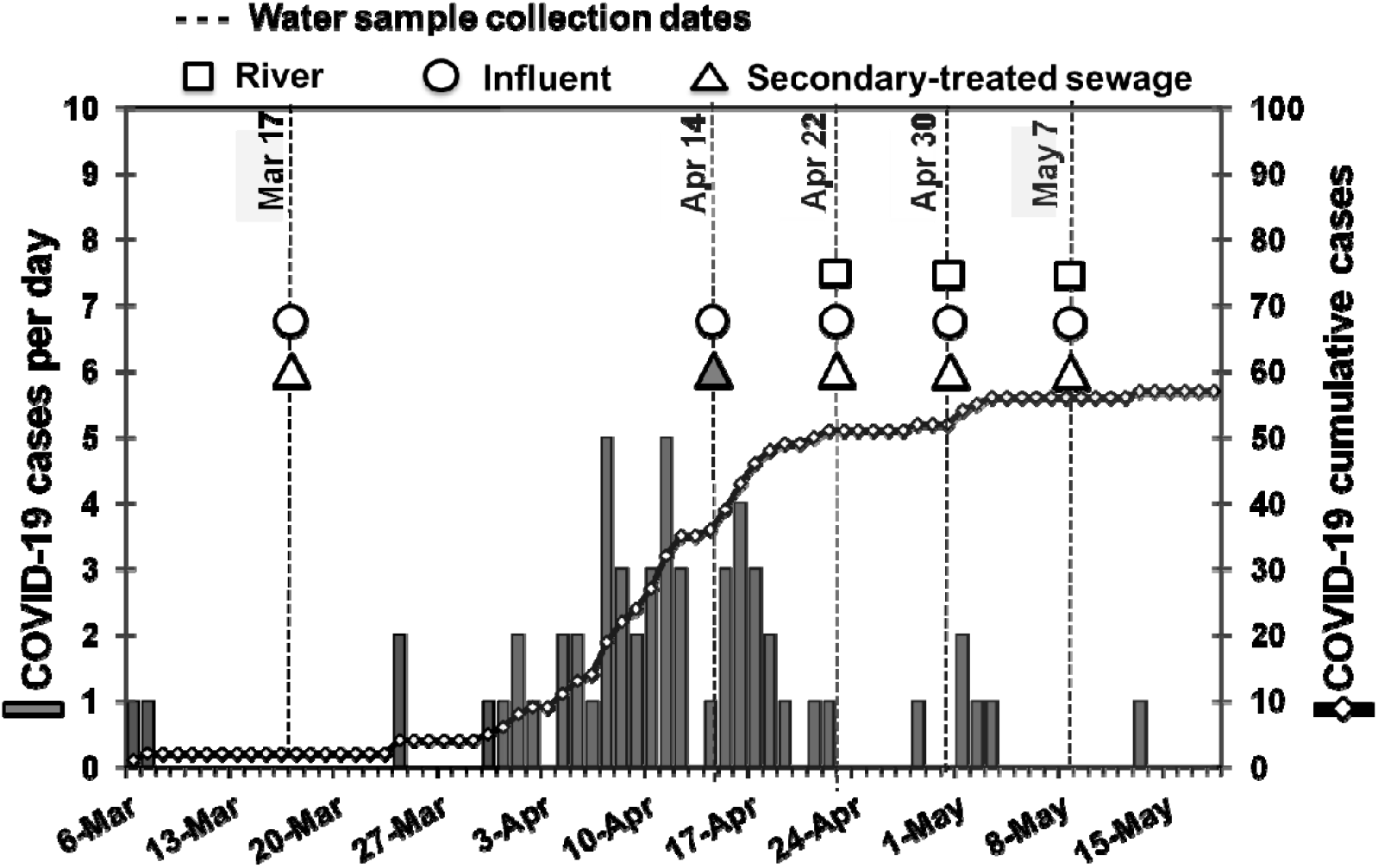
COVID-19 cases and SARS-CoV-2 RNA detection in wastewater and river water in Yamanashi Prefecture, Japan. Squares, circles, and triangles denote sampling dates of river water, influent and secondary-treated sewage samples, respectively. A closed triangle denotes SARS-CoV-2 RNA detection, grey bars denote new COVID-19 cases on each day, and black thread line with white diamonds denotes COVID-19 cumulative cases in Yamanashi Prefecture.

The threshold cycle (Ct) of the positive secondary-treated wastewater sample was 39.96, which corresponds to 2.4 × 10^3^ copies/L in the original water sample. This result was further confirmed to check for any accidental contamination of the sample with the gBlocks positive control by using a VIC-labeled check probe (5’-VIC-AGCTAGCGCATTGGATCTCG-NFQ-MGB-3’) (Shirato et al., 2020), where the VIC fluorescent signal was detected from the positive control containing the check probe sequence but not detected from the positive sample. Furthermore, the positive qPCR mixture was purified using a QIAquick PCR Purification Kit (Qiagen), followed by direct nucleotide sequencing, where forward and reverse sequences were closely identical to SARS-CoV-2 strains available in GenBank database (data not shown). In addition, the obtained nucleotide sequences did not match with the check probe sequence.

The cumulative COVID-19 cases in Yamanashi Prefecture was 36 on April 14, 2020, when a positive signal from the secondary-treated wastewater sample was obtained (Fig. 1).

## 4. Discussion

In the current study, two different methods (EMV and adsorption-direct RNA extraction methods) were applied for concentration-RNA extraction of SARS-CoV-2. For this purpose, PMMoV was selected as an indicator virus, considering its extremely high abundance in wastewater (Kitajima et al., 2018). The EMV method outperformed the adsorption-direct RNA extraction method with approx. 2.4-log_10_ higher observed concentrations of indigenous PMMoV RNA in the samples. The adsorption-direct RNA extraction method was expected to work well as the recovery of murine hepatitis virus (MHV) was high in a recent comparison study, although HA filter (0.45-µm pore-size, 47-mm diameter electronegative membranes) was used (Ahmed et al., 2020b), while AA filter (0.8-µm pore-size, 90-mm diameter electronegative membranes) was used in the current study. However, unlike SARS-CoV-2, PMMoV is a non-enveloped RNA virus. Besides, PMMoV is negatively charged, whereas coronaviruses are positively charged at neutral pH. Thus, the recovery of an enveloped surrogate virus with a similar structure to SARS-CoV-2, such as MHV, transmissible gastroenteritis virus, and *Pseudomonas* phage Φ6, should be evaluated in future studies (Kitajima et al., 2020).

Of the ten wastewater samples tested, SARS-CoV-2 RNA was detected only in 20% (1/5) of secondary-treated wastewater samples by N_Sarbeco qPCR assay following the EMV method, which had been collected on April 14, 2020. SARS-CoV-2 RNA has been detected in wastewater in Australia by N_Sarbeco assay (Ahmed et al., 2020),, in the Netherlands by CDC-N1, -N2, -N3, and E_Sarbeco assays (Medema et al., 2020), and in Spain by CDC-N1, -N2, and -N3 assays (Randazzo et al., 2020).).

None of the river water samples tested positive for SARS-CoV-2 RNA using six qPCR/nested PCR assays, following the EMV and the adsorption-direct RNA extraction methods. The possible reason for this could be a low prevalence of COVID-19 infections in the studied region.

In the present study, despite the detection of SARS-CoV-2 RNA in the secondary-treated wastewater sample, it was not detected in any of the influent samples tested following the same procedure. This discrepancy in the results could be due to the difference in the initial volume of water samples used for concentration. The volume of secondary-treated wastewater samples (5,000 mL) filtered was 25 times greater than that of the influent samples (200 mL), which leads to higher LOD for the influent samples (4.0 × 10^3^−8.2 × 10^4^ copies/L) as compared to LOD for the secondary-treated wastewater samples (1.4 × 10^2^−2.5 × 10^3^ copies/L). In addition, hydraulic retention time for the wastewater treatment process was not considered while collecting the samples (i.e., both influent and secondary-treated wastewater were collected almost at the same time). The SARS-CoV-2 RNA concentration in the positive secondary-treated wastewater sample was two orders of magnitude lower (2.4 × 10^3^ copies/L; Ct of 39.96 in only one of two PCR wells) than that reported in a previous study in Spain (2.5 × 10^5^ copies/L) (Randazzo et al., 2020). Randazzo et al. (2020) reported comparable level of SARS-CoV-2 RNA concentrations between influent and treated wastewater samples in Spain (both approx. 2.5 × 10^5^ copies/L). Thus, there is a necessity to perform further studies on reduction of SARS-CoV-2 virus in a WWTP.

Out of five sampling dates of this study, SARS-CoV-2 was not detected in four dates (March 17, April 22, 30, and May 7) and detected only on April 14. Before March 17, the reported cumulative cases of COVID-19 in Yamanashi Prefecture was only 2. As of April 14, when SARS-CoV-2 RNA was detected in the current study, 36 cumulative cases had been reported, corresponding to 4.4 cumulative cases per 100,000 inhabitants. The curve of cumulative cases started to rise after March 30. The highest daily new reported cases (5 cases/day) were recorded on April 7 and 11 after which the daily new cases reduced noticeably. The curve of cumulative COVID-19 cases started to flatten after April 16. Thus, we could observe the correspondence between the time of detection of the virus in our samples and the time of the highest peak in the number of daily cases of COVID-19 infection in the prefecture.

As of June 1, Yamanashi Prefecture has been one of the prefectures with relatively low COVID-19 prevalence in Japan. Lower infection prevalence could be responsible for the lower detection ratio of SARS-CoV-2 RNA in wastewater samples in this study. Nevertheless, detection of SARS-CoV-2 RNA (even in low concentration) that corresponded to the peak of daily new cases of infection provided an assurance that even in the areas of low prevalence, WBE can be used as tracking or warning tools for monitoring the status of COVID-19 prevalence in a community. SARS-CoV-2 RNA detection in wastewater has been reported from a country with a low prevalence of COVID-19 cases, and even prior to COVID-19 cases had been reported (Randazzo et al., 2020). WBE could be an effective and economical tool to monitor the status of SARS-CoV-2 circulating within a community to reduce the risk of future outbreaks (Ahmed et al., 2020; Hart and Halden, 2020; Kitajima et al., 2020; Medema et al., 2020; Orive et al., 2020; Xagoraraki and O’Brien, 2020). More in-depth investigations on the spread of the virus through wastewater and on ascertaining the role of water and sanitation interventions to prevent waterborne transmission have been suggested (Heller et al., 2020; Nunez-Delgado 2020).

Considering the low concentrations of SARS-CoV-2 in wastewater, further studies to explore a more effective concentration-RNA extraction method is recommended. Detection of SARS-CoV-2 RNA in wastewater can be affected by many factors, including concentration-RNA extraction methods, RT-qPCR assay, and the prevalence of COVID-19 infections in the community. Besides these determinants, some country-specific factors, such as differences in per-capita water use or the sewer systems channeling domestic sewage and stormwater to WWTPs (e.g., combined or separate sewer), might influence the concentration of viral RNA in wastewater reaching a WWTP.

Despite the successful detection of SARS-CoV-2 RNA in secondary-treated wastewater, it is uncertain if the detected viral RNA was infectious at the time of sampling. Besides, secondary-treated wastewater is further chlorinated before being discharged to an open water body. Coronaviruses are generally known to be sensitive to chlorine and inactivated relatively faster in water compared to non-enveloped viruses, such as PMMoV (La Rosa et al., 2020). Wang et al. (2005) reported a complete inactivation of SARS-CoV in wastewater with chlorine (10 mg/L for 10 min; free residual chlorine, 0.4 mg/L) or chlorine dioxide (40 mg/L for 30 min; free residual chlorine, 2.19 mg/L). Chlorine-based disinfectants, such as household bleach, chloroxylenol, chlorhexidine, and benzalkonium chloride, were found effective for inactivation of SARS-CoV-2 (Chin et al., 2020). Furthermore, comparable levels of reductions of *E. coli* and PMMoV between the current study and a previous study (Tandukar et al., 2020) supported that these pathogens are removed substantially at the WWTP and the treatment systems are functioning appropriately. However, unfortunately, in the current study, SARS-CoV-2 RNA reduction could not be calculated because it was not detected in the influent samples.

There are other cities in Japan that have low reported cases of COVID-19. Because asymptomatic cases are less likely to be reported to public health officials, WBE could be an effective disease surveillance tool for such cities. In addition, WBE could serve as a large scale, population-wide surveillance in near real-time (Sims and Ksprzyk-Hordern, 2020), especially in resource-limited regions and to alert emergency response teams for preparedness. A unified platform, such as ‘COVID-19 WBE Collaborative’ consisting of scientists from multiple discipline that aims to facilitate timely and high-impact WBE studies for public benefit, is required to fight against the ongoing COVID-19 pandemic.

## 5. Conclusions

- SARS-CoV-2 RNA was detected in one of five secondary-treated wastewater samples collected from a WWTP (2.4 × 10^3^ copies/L) by N_Sarbeco qPCR assay following the EMV method, which serves as the first case of detection in wastewater in Japan.
- SARS-CoV-2 RNA was not detected in any of the five influent and three river water samples tested using four qPCR (N_Sarbeco, NIID_2019-nCOV_N, and CDC-N1 and - N2) and two nested PCR (ORF1a and S protein) assays.
- Even when the number of reported COVID-19 cases was low (4.4 cumulative cases per 100,000 inhabitants), SARS-CoV-2 RNA was detected in a secondary-treated wastewater sample when the weekly reported cases in the community were high.
- Based on the observed concentrations of indigenous PMMoV RNA, the EMV method followed by RNA extraction using the QIAamp Viral RNA Mini Kit performed better than the adsorption-direct RNA extraction method followed by RNA extraction using the RNeasy PowerWater Kit, suggesting the applicability of the EMV method for detection of SARS-CoV-2 RNA in wastewater.

## Data Availability

All data are shown in the manuscript.

## Authorship contribution statement

**Eiji Haramoto:** Conceptualization, Funding acquisition, Investigation, Resources, Supervision, Writing-reviewing and editing, Formal analysis. **Bikash Malla:** Writing-original draft, Formal analysis, Laboratory work. **Ocean Thakali:** Reviewing, Formal analysis, Laboratory work. **Masaaki Kitajima:** Conceptualization, Investigation, Resources, Writing-reviewing and editing, Formal analysis.

## Acknowledgements

This study was partially supported by the Japan Society for the Promotion of Science (JSPS) through Grant-in-Aid for Scientific Research (B) (grant number JP20H02284). The authors acknowledge the staffs of the WWTP for their support and permission on wastewater sampling.

